# Improving newborn health with family-centered, early postnatal care: a quasi-experimental study

**DOI:** 10.1101/2022.10.11.22280956

**Authors:** Seema Murthy, Shirley Du Yan, Shahed Alam, Amit Kumar, Arjun Rangarajan, Meenal Sawant, Huma Sulaiman, Bhanu Pratap Yadav, Tanmay Singh Pathani, Anand Kumar H.G., Sareen Kak, Vinayaka A M, Baljit Kaur, Rajkumar N, Archana Mishra, Edith Elliott, Megan Marx Delaney, Katherine E.A. Semrau

## Abstract

Despite the recent decline, neonatal mortality rates (NMR) remain high in India. Family members are often responsible for the postpartum care of newborns and mothers. Yet, low health literacy and varied beliefs can lead to poor neonatal health outcomes. Postpartum education for family caregivers can improve the adoption of evidence-based newborn care and health outcomes. The Care Companion Program (CCP) is a hospital-based, pre-discharge health training session where nurses teach key healthy behaviors and help mothers and family members learn skills and practice in the hospital. Here, we assessed the impact of CCP on NMR. We conducted a quasi-experimental study to assess the effect of the CCP sessions on mortality outcomes among families seeking care in 28 public tertiary facilities, across 4 Indian states. Neonatal mortality outcomes were reported post-discharge, collected via phone surveys at four weeks of age of baby, between October 2018 to February 2020. Risk ratios (RR), adjusting for hospital-level clustering, were calculated by comparing mortality rates before and after CCP implementation. A total of 46,428 families participated in the pre-intervention group and 87,305 in the post-intervention group; 76% of families participated in the phone survey. The crude NMR was 33.64 deaths per 1000 live births (RR=0.82, 95% CI: 0.76, 0.87). After accounting for hospital-clustering, the NMR was 41.3 (adjusted RR=0.81, 95% CI: 0.71, 0.93). There may be a substantial benefit to family-centered education in the early postnatal period to reduce neonatal mortality.

## Introduction

Neonatal mortality disproportionately affects families in low-to-middle income countries (LMICs) (1,2). With 21.7 deaths per 1000 live births, India has the third-highest Neonatal Mortality rate (NMR) (3). India’s NMR has declined sharply from 1990 (57.4 deaths) (4). However, current strategies to improve neonatal survival include increasing institutional deliveries, increasing coverage of care, creating structured infrastructure across communities for the care of at-risk babies, and improving healthcare access have reached a ceiling effect (5). To further improve neonatal survival, there is a need for different approaches, and more innovative targeted interventions (6,7).

Most deaths can be prevented by providing Continuum of Care (CoC) in a structured way from pregnancy to the post-delivery period (8). Interventions that take a lifecycle approach addressing the needs of the woman before, during, and after her pregnancy, as well as infant and childcare, are required. Although institutional deliveries have nearly doubled in India over the past decade (9), insufficient follow-up visits create gaps in postnatal care. According to the National Family Health Survey (NFHS-4), only 39% of pregnant women in India had completed CoC between 2015 and 2016, including the utilization of four major aspects: 4 + antenatal care, institutional delivery, postnatal care, and immunization (8). In LMICS, only 58% of babies complete the postnatal visit within two days after birth (10). Besides insufficient follow-up care, caregivers often lack knowledge of correct practices for providing care and/ or identifying critical situations where additional and immediate care is required for the newborn (10). It is possible that lack of required follow-up visits and absence of knowledge for adequate postnatal care can have a combined effect on the baby and/or mother’s health increasing the risk of mortality and morbidity.

Although certain healthcare interventions can only be provided by skilled healthcare personnel, there are many practices that caregivers can and should do on an ongoing basis, but the caregivers need to be first equipped through training to do so (10). Postnatal training is an important strategy to increase the skills of caregivers and families. With the increase in institutional deliveries, hospital stays provide a window of opportunity to equip caregivers with the necessary awareness and skills. A hospital-based program that can systematically train caregivers, could potentially address this gap in caregiver knowledge, leading to improved care practices and health outcomes.

### Program description

The Companion Program (CCP) trains families on essential maternal and newborn care during their hospital stay using a novel approach. Noora Health, a non-profit organization, supported the design of CCP, which is being implemented as a public-private partnership across district hospitals in four states of India (Karnataka, Madhya Pradesh, Maharashtra, and Punjab) (11). The program has an evidence-based curriculum for postnatal counseling created using a human-centered design process to promote behavior change. It includes behaviors for improving neonatal and maternal health (12). The behaviors taught include early and exclusive breastfeeding; hygienic cord care; hand hygiene; skin-to-skin care (kangaroo mother care); burping; healthy diet for the mother; the importance of follow-up and immunizations; and recognition of all danger signals with respiratory infections, diarrhea, and jaundice being covered in more detail. Information is distributed using visual aids and materials tailored to the cultural practices of local communities and patient populations (12). Through the CCP, nurses and health educators receive training on health behavior change and skills to effectively communicate health information by engaging their audiences. They are provided teaching aids that include videos played on television monitors in the postnatal wards, flipcharts, demonstration tools (e.g. dolls) used for role-playing, and hand-outs. Using these varied materials, nurses and health educators conduct group classes for families in postnatal wards. Additionally, families receive post-discharge follow-up through a dedicated WhatsApp-based platform, which reiterates key behavior change messages, and allows for two-way communication, providing families the opportunity to ask questions and receive guidance. Initial findings showed promising results for increased behavior change, and reduction in preventable complications and readmissions (13).

An ongoing large evaluation study is assessing the impact of this intervention on knowledge, practice, and outcomes among families seeking care in 28 facilities, across 4 Indian states. The data collection for the main study was temporarily paused due to the COVID-19 pandemic. Here, we are reporting the findings from the interim analysis assessing the early impact of the CCP on neonatal mortality.

## Methods

### Study procedure

A quasi-experimental study design was used to assess the impact of CCP on neonatal outcomes. Data collected from October 2018 to Feb 2020 was analyzed for mortality outcomes in this interim analysis. Among the 28 study hospitals, 24 district-level hospitals (DHs) were selected randomly from 91 high and low delivery-load hospitals in 3 Indian states, Karnataka (KA), Punjab (PB), and Madhya Pradesh (MP), in which CCP would be implemented shortly. In Maharashtra (MH), four government medical college hospitals were also included.

Trained investigators collected contact details of the mothers in the hospital, after delivering the baby, and just before discharge. Mother’s death or neonatal death before discharge were excluded from the study. Mothers who were less than 18 years of age or did not give consent were excluded from the study. At the end of the neonatal period, the study staff called the consenting and eligible mothers to learn about newborn care behaviors. Those who were not reached or those who refused the interview were marked as incomplete. The investigators did not complete follow-up questions about newborn behaviors or demographics if either the mother or newborn death was reported at the time of call, due to ethical concerns of continuing the interview. As a result, no adjustments are possible based on demographic data.

The pre-intervention data collected before the CCP launch date at each facility was compared to post-intervention data, which was collected after the launch date of the program at that facility. We measure the effect of the program with an intention-to-treat approach, irrespective of whether individuals in the intervention facilities were trained or not. No time elapsed between when the program started, and data collection began. We compared the post-discharge mortality for neonates in the two groups. Maternal mortality data were also collected, but are presented in Appendix A.

### Sample size

This sample size calculation at the time of designing the study accounted only for behavior change. Hence, a post hoc sample size calculation to estimate the power of the study to detect NMR and MMR estimated in the study was performed using R version 4.1. With the current sample size, the study has at least 98% power to detect the estimated difference in NMR of 8.7 deaths per 1000 live births between the pre- and post-CCP session groups. A post-hoc power calculation was done that was sufficient to detect the differences.

### Analysis plan

We used logistic regression models to assess the impact of the intervention (pre versus post) on mortality. Our dataset was limited to those who reported outcomes during a follow-up call. There were 2 cases for whom information on any of the survey-related information was not collected. Hence, they were dropped from the analysis. In all subsequent analyses, cases were analyzed based on the availability of the data. The regression models were adjusted for hospital-level clustering. We reported hospital-cluster adjusted risk ratios and risk differences. All analyses were done using STATA 16 (14).

The regression model was:

Log [P(Mortality in mother/Baby | Group)/(1-P(Mortality in mother/Baby| Group))]= B0+B1*Group +error B0, B1 are hospital-level cluster adjusted estimates

### Ethical review

We received IRB approval for primary data collection from ACE Ethics Committee and SPECT. The primary data with identifier details rested with the core team in India. The deidentified data was shared with collaborators for analysis. For this secondary analysis (DCGI Reg. No. ECR/141/Indt/KA/2013), Harvard University Institutional Review Board (Protocol # IRB19-1140) deemed the study exempt. All participants underwent an oral consent process, which was audio recorded.

## RESULTS

### Sample flow

We collected phone numbers from a total of 133,733 families in the hospital, with 46,428 from the Pre group and 87,305 from the Post group (Fig 1). We successfully contacted and ascertained mortality outcomes in 33,599 (72.4% response rate) of the pre-intervention group and 60,078 (68.8% response rate) of the post-intervention group (overall response rate of 70.6%). The primary reasons for non-response in both the groups were due to the phone line not even being connected, invalid/ wrong number, and never picked/unavailable. Fewer people refused, spoke a different language than the data collector, or were ineligible as per the exclusion criteria. Under ineligibility, the most frequent criterion was that the baby was delivered to a different facility and then shifted to the current hospital.

**Fig 1.**
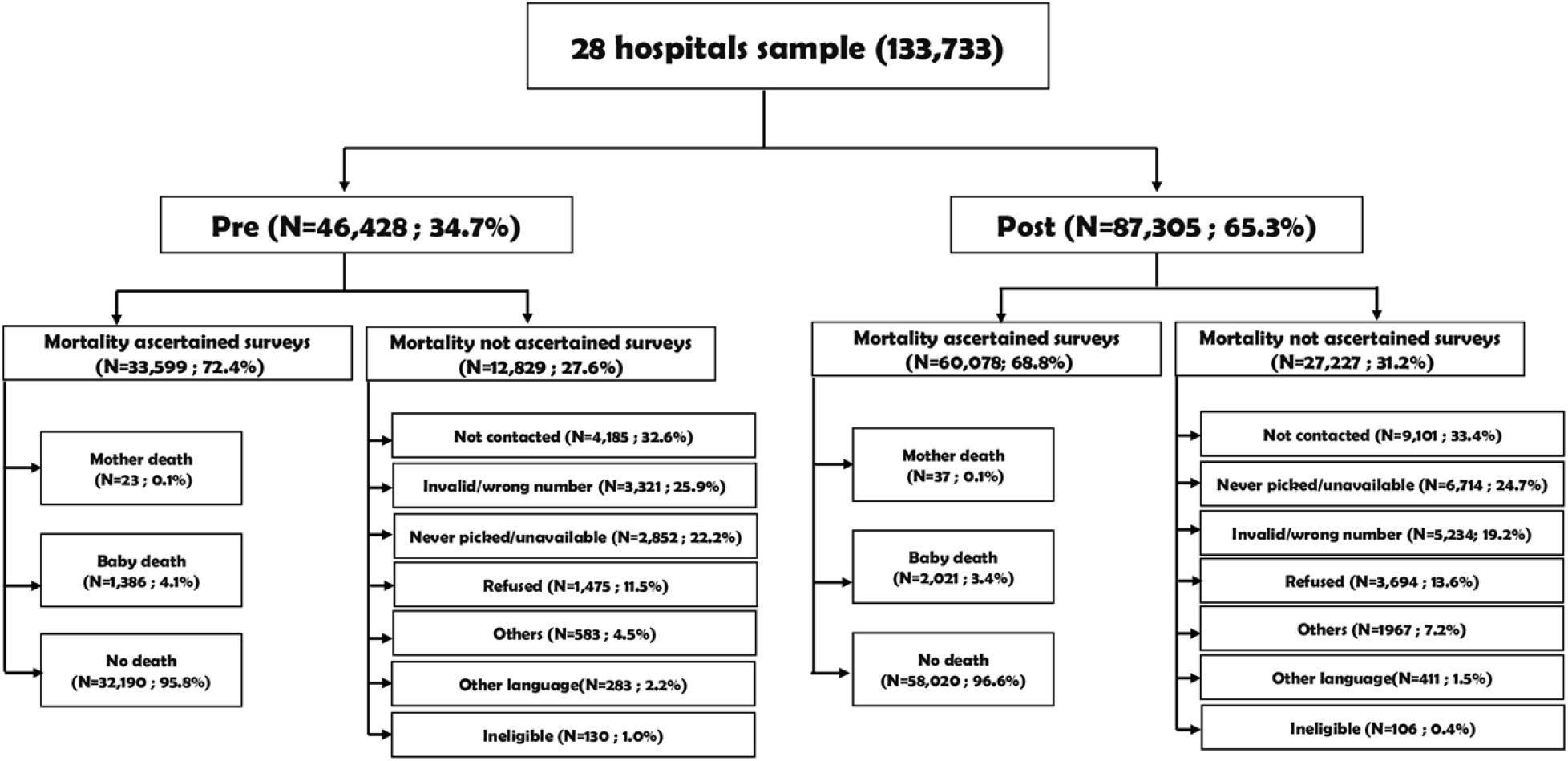
Flowchart and Sample Size.

### Description of the cohort

Among the interviewed population that did not experience a death, we describe the study cohort pre- and post-intervention (Table 1). Although this does not include the details of the deaths it serves to describe the population cohort. Overall, there were no differences of more than 10% between pre- and post-intervention groups. There were more younger mothers (age <=25 years) in the pre-group (72.2%) as compared to the post-implementation group (66.4%). Education was slightly different, with the biggest difference in the people receiving 1-8 years of education in the pre-group being 4.6% less in the post-group. About 4% more people studied 12 years in the post group. More people (about 6%) had C-section deliveries in the post group. The number of babies delivered, the length of stay in the hospital, and whether this was the mother’s first baby were the same between pre- and post-groups. All socio-demographic comparison findings between pre- and post-groups were statistically significant (p<0.05) except for primiparas.

**Table 1.**
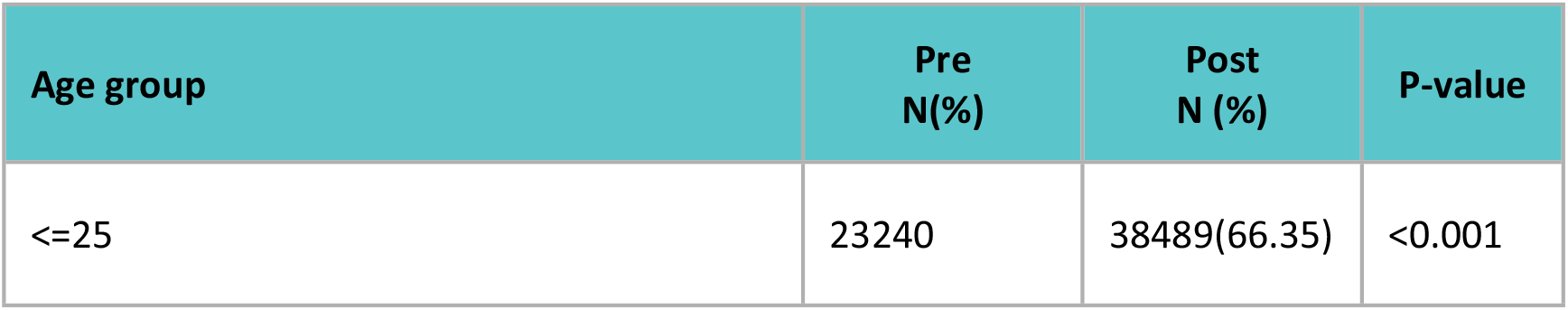
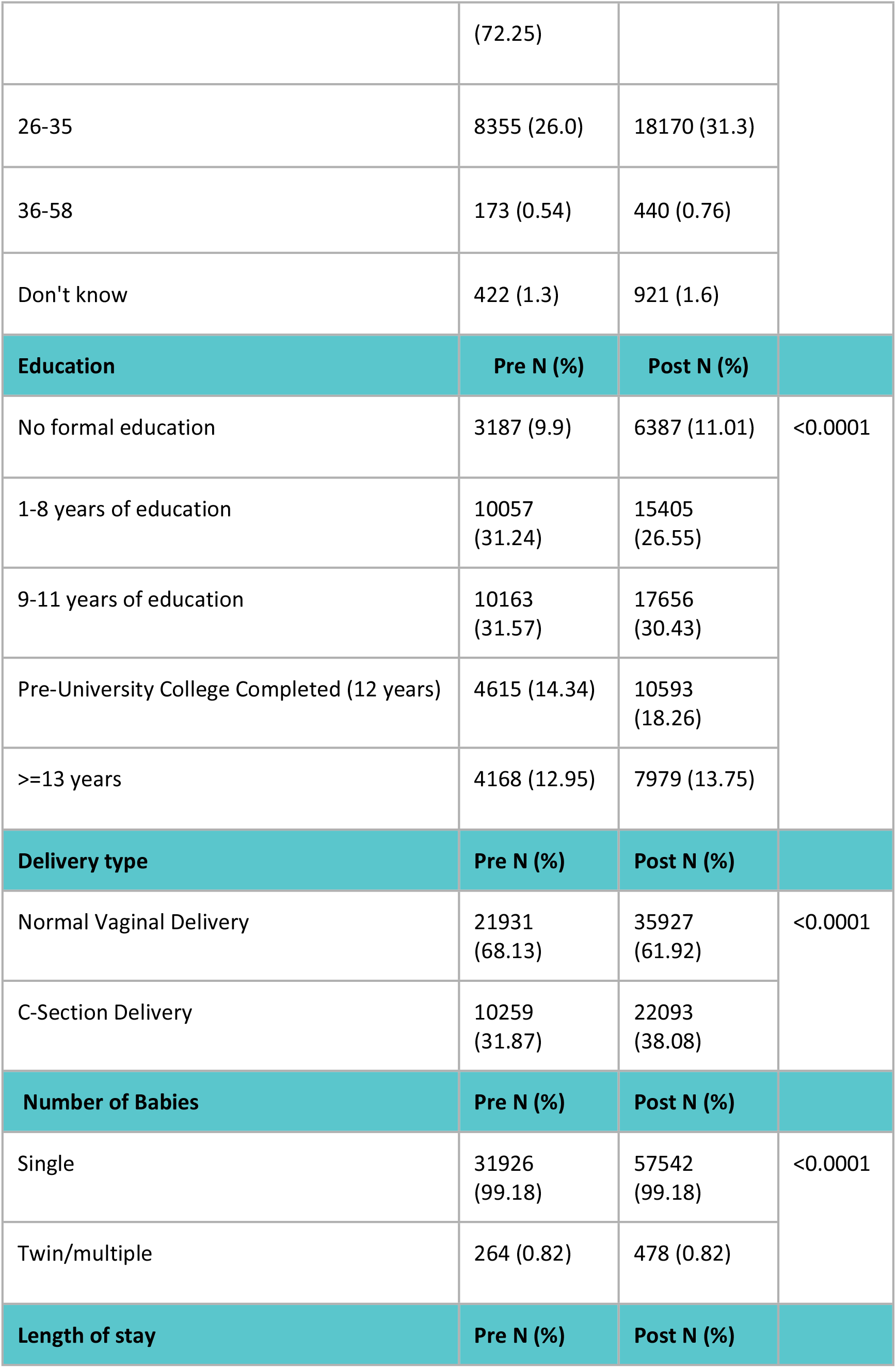
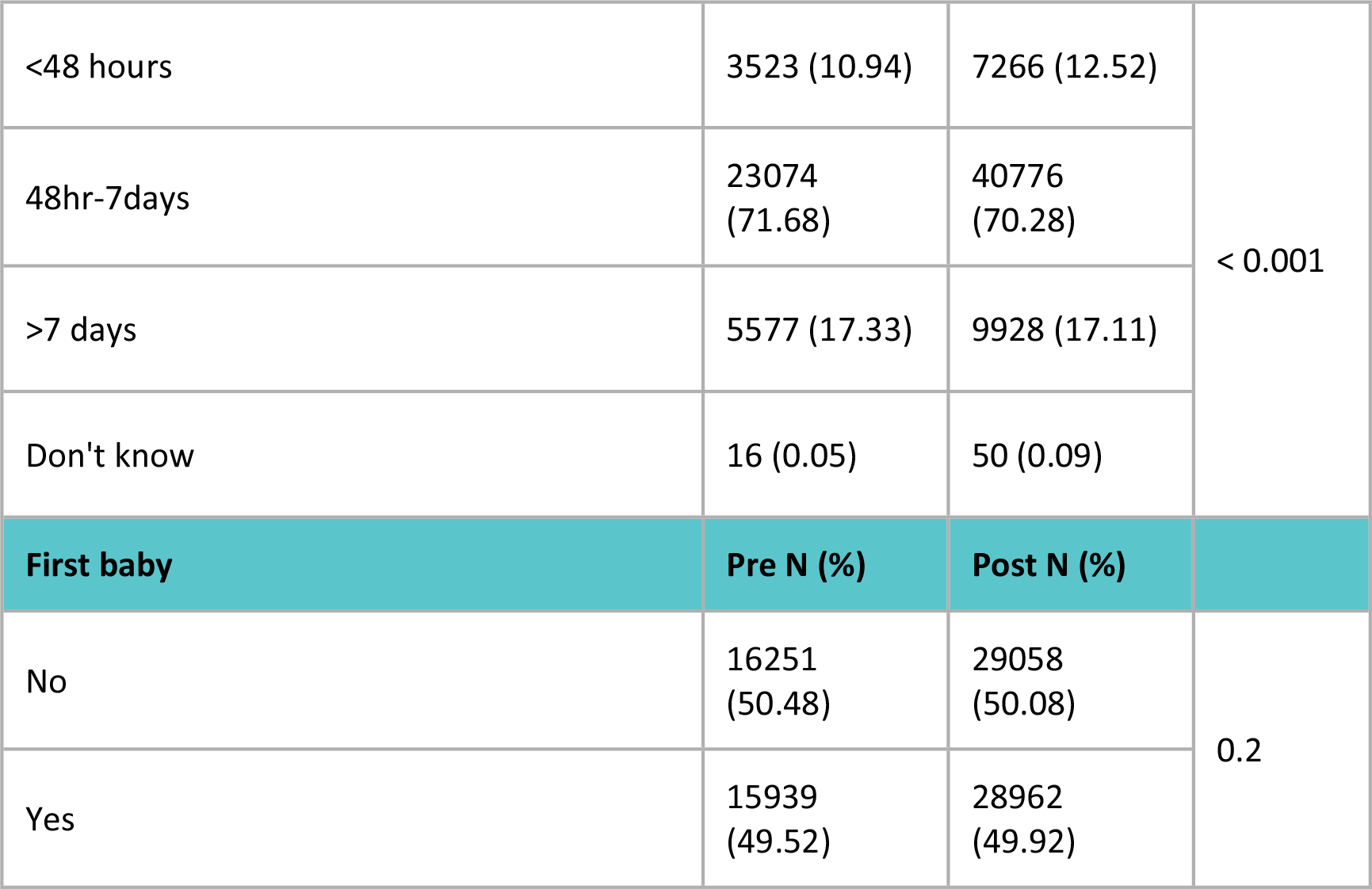
Socio-demographic Characteristics of Respondents.

| Age group | Pre N(%) | Post N (%) | P-value |
| --- | --- | --- | --- |
| $\leq 25$ | 23240 | 38489(66.35) | <0.001 |
|  | (72.25) |  |  |
| 26-35 | 8355 (26.0) | 18170 (31.3) |  |
| 36-58 | 173 (0.54) | 440 (0.76) |  |
| Don't know | 422 (1.3) | 921 (1.6) |  |
| **Education** | **Pre N (%)** | **Post N (%)** |  |
| No formal education | 3187 (9.9) | 6387 (11.01) | <0.0001 |
| 1-8 years of education | 10057 (31.24) | 15405 (26.55) |  |
| 9-11 years of education | 10163 (31.57) | 17656 (30.43) |  |
| Pre-University College Completed (12 years) | 4615 (14.34) | 10593 (18.26) |  |
| >=13 years | 4168 (12.95) | 7979 (13.75) |  |
| **Delivery type** | **Pre N (%)** | **Post N (%)** |  |
| Normal Vaginal Delivery | 21931 (68.13) | 35927 (61.92) | <0.0001 |
| C-Section Delivery | 10259 (31.87) | 22093 (38.08) |  |
| **Number of Babies** | **Pre N (%)** | **Post N (%)** |  |
| Single | 31926 (99.18) | 57542 (99.18) | <0.0001 |
| Twin/multiple | 264 (0.82) | 478 (0.82) |  |
| **Length of stay** | **Pre N (%)** | **Post N (%)** |  |
| <48 hours | 3523 (10.94) | 7266 (12.52) | < 0.001 |
| 48hr-7days | 23074<br>(71.68) | 40776<br>(70.28) |  |
| >7 days | 5577 (17.33) | 9928 (17.11) |  |
| Don't know | 16 (0.05) | 50 (0.09) |  |
| **First baby** | **Pre N (%)** | **Post N (%)** |  |
| No | 16251<br>(50.48) | 29058<br>(50.08) | 0.2 |
| Yes | 15939<br>(49.52) | 28962<br>(49.92) |  |

#### Mortality

The unadjusted Risk Ratio (RR) for neonatal mortality in the post versus pre group was 0.82 (95% CI:0.76, 0.87). Cluster-adjustment did not alter the RR (0.81, 95% CI: 0.71, 0.93) (Table 2 and 3).

**Table 2.** Unadjusted Estimates of Neonatal Mortality Rate.

| Indicator | Pre*<br>N=33599 |  | Post<sup>†</sup><br>N=60078 |  | Risk Ratio | 95% CI<sup>‡</sup> |
| --- | --- | --- | --- | --- | --- | --- |
|  | Deaths (n) | Estimate | Deaths (n) | Estimate |  |  |
| NMR<br>(per 1000 live<br>births) | 1386 | 41.26 | 2021 | 33.64 | 0.82 | 0.76, 0.87 |

\*Pre, Standard of Care
<sup>†</sup> Post, Care Companion Program
<sup>‡</sup> CI confidence Interval

**Table 3.**
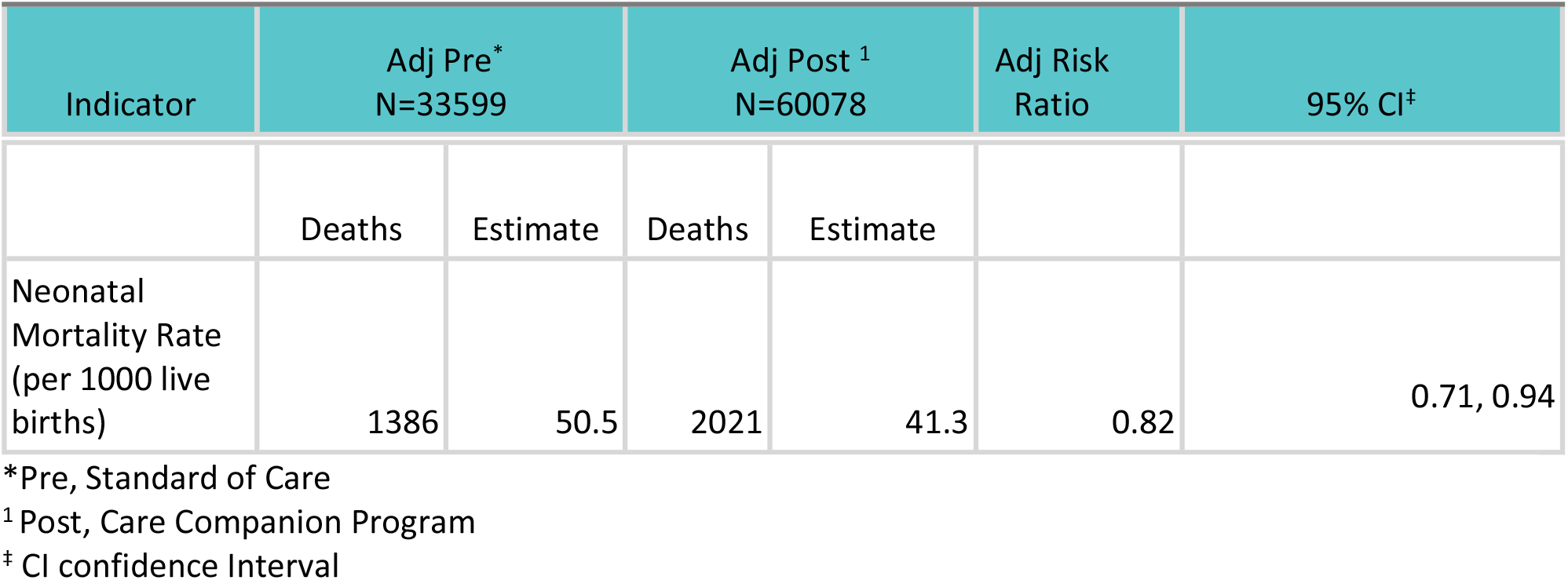
Neonatal Mortality Rate and Post-discharge 28 day.

The trendline between pre- and post-implementation of CCP compared the larger trendline of the World Bank (WB) reported NMR in India (Fig 2). The green line indicates when CCP began implementation across study sites. The trendline of NMR decrease is steeper than the World Bank NMR (15), showing the association of CCP towards reducing NMR.

**Fig 2.**
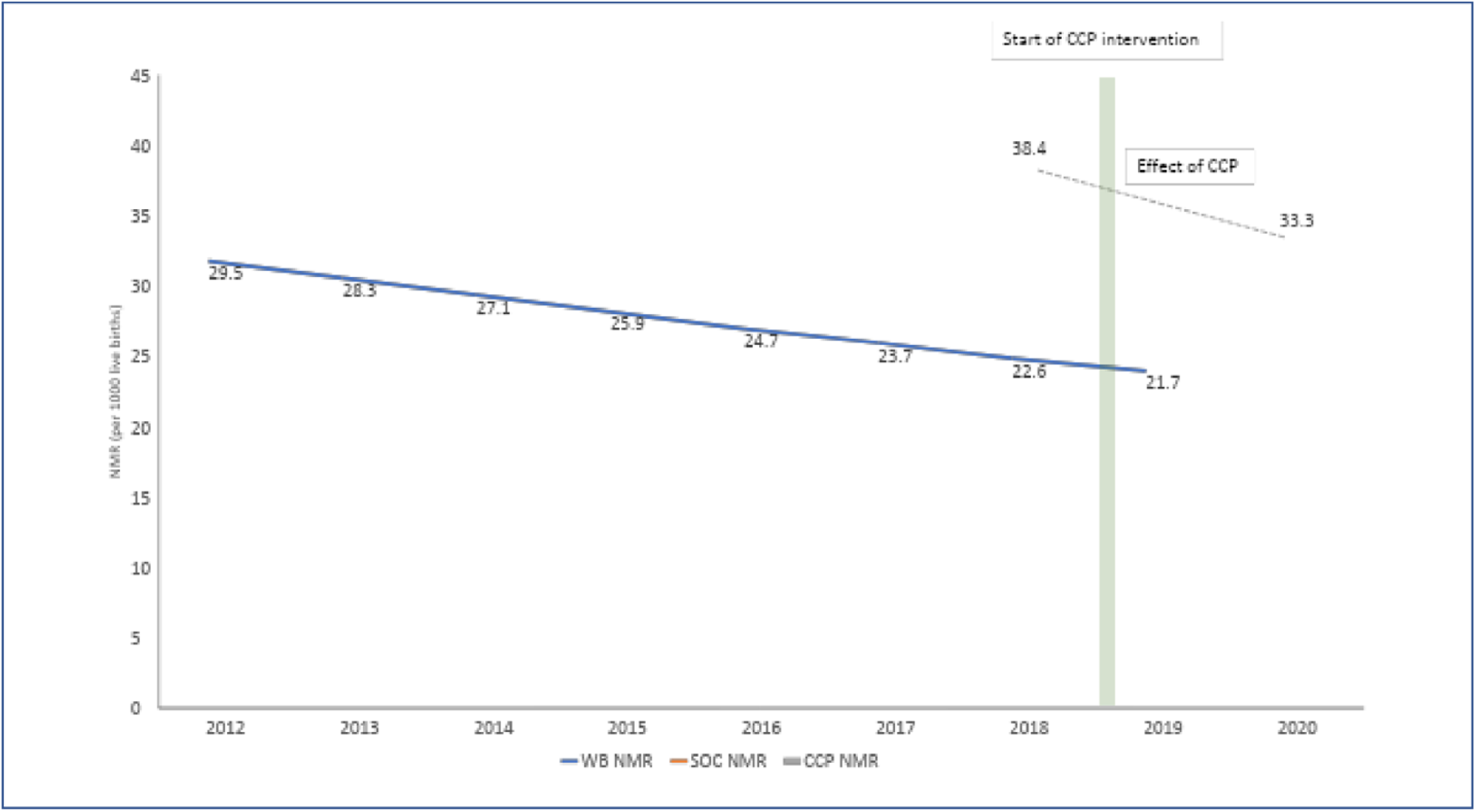
Associated Impact of CCP on Neonatal Mortality. WB: World Bank SoC: Standard of Care, pre CCP implementation CCP: Care companion Program

Maternal mortality was not significant. In Appendix Table A, neonatal mortality adjusted for state and hospital-level clustering, and crude model by the state are reported. Appendix Table B outlines NMR rates as reported NFHS-4 and -5; Table C outlines dates for when pre and post-data collection began, and Table D reports estimates of maternal mortality.

## DISCUSSION

In this large cohort, CCP was associated with a reduced NMR compared to the pre-implementation period. CCP promotes behavior change at the family level and reinforces positive behaviors to improve the health of the baby. It also helps families identify early warning signs and encourage appropriate health-seeking behavior. We hypothesize that all of this leads to an associated reduction in neonatal mortality among families who receive CCP training. Our results show a viable strategy for newborn outcomes following birth.

Due to a shortage of staff and a lack of knowledge among parents about correct newborn care practices, Family-centered Care (FCC) is becoming a need of time for postnatal care (16). Training physicians and nurses on delivering important medical information to patients as well as educating patients and caregivers are two key components of FCC (16). FCC-based interventions have resulted in positive health outcomes in several conditions (13,17–19). Promotion of antenatal care and maternal health education is given to mothers and caregivers to improve key behaviors resulting in positive health outcomes (20–22). Hospital-based FCC interventions focusing on awareness and improving the skills of parents and caregivers can be helpful in improving key behaviors and postnatal outcomes (13). Similarly, FCC-based community interventions include sociocultural appropriate behavior modification techniques such as delivering health education and information to parents and caregivers, training birth attendants on identifying early danger signs, and training community health workers as master trainers to further train the family members on risky maternal and newborn care practices were expected to reduce the neonatal mortality and morbidity burden on health systems (22,23). Reduced NMR in the post-CCP group found in this study supports this evidence.

The NMR observed in our study (pre: 50.3/1000 post: 41.3/1000) is much higher than the national average in the same time frame (24.9/1000) (24). This difference may be explained by the study population and CCP program at secondary and tertiary levels where Special Newborn Care Unit babies were also included. Focusing the family health education on a hospital setting with more high-risk babies likely had a greater impact on mortality.

The change in mortality rates over time is varied in time and different populations. An average may be an oversimplification for looking at secular trends. We considered if the lower mortality seen in the post-implementation period may have been driven by secular trends or concurrent Maternal and Newborn Health (MNH) initiatives. Historically, NMR reduced by 9 per 1000 births (39 to 30 per 1000 births), over a much longer 10-year time frame, between NFHS-3 and 4 (2005–06 and 2015–16 respectively) (25). In the 1.5 years of program implementation, this translates to 9.2 per 1000 births lower and an 18% reduction in adjusted estimates (Fig 2). Thus, the secular trend explains only a part of the difference we have seen. We did not see any new Maternal and Child Health (MNCH) initiatives occur in all intervention facilities in the 1.5 years between the pre- and post-data collection. However, programs like LaQshya and non-health-related programs which took place in a few facilities could have contributed to the reduction. We believe that a large part of the difference seen is due to the direct training offered by CCP and the involvement of the whole family in behavior change.

The strengths of our study are the large sample size and representative data from multiple government facilities across 4 states. We adjusted for the cluster effect of the site using a logistic regression model to get a range around the point estimate. We have used a robust study design including a comparison group from the same hospitals, similar populations seeking care, and similar settings reducing the variability. The demographics of the cohort for the completed surveys (without death) show statistically significant differences in the pre-and post-groups. There were typically older mothers, with more C-sections in the Post group, which may have a higher risk of mortality. We were unable to adjust for these differences in the Pre and Post groups, yet this group showed that mortality was reduced despite the Post group being at slightly higher risk.

There are four key limitations of our data. First, we could not measure the outcomes in the non-responders. We have compensated for this by calculating the NMR by including a denominator where we know the outcomes. Second, state-level estimates were not found to be significant, whereas the pooled estimates were. The tables for these analyses can be found in the Appendix in Table A. Third, for ethical reasons, we did not proceed with the survey when death was reported and thus we have limited information to adjust for potential confounders, the cause, and timing of death post-discharge. Fourth, as an implementation research study design, we did not randomize individuals or clusters to receive the intervention and all data are self-reported. The study was not originally powered to detect changes in mortality.

## CONCLUSION

Postnatal training for caregiver skills could supplement health-system existing efforts to reduce neonatal mortality. The estimated trend toward reduction in neonatal mortality over this short time span during CCP implementation is encouraging. Further studies should collect additional demographic details around deaths to understand at-risk populations. Further death reviews ascertaining the cause and timing of death as well as further exploring any causal pathways such as knowledge and practice of caregiving behaviors can help elucidate potential mechanisms involved in reducing mortality. Family-centered approaches to healthcare can help ensure proper patient care extends beyond hospitals into homes and lead to improved health outcomes. Noora Health plans to expand CCP’s footprint and evaluate the impact of caregiver education across condition areas and geographies.

## Data Availability

Complete data will be available upon request

## ACKNOWLEDGEMENTS

We would like to acknowledge the contributions of the larger Noora Health Implementation and Research teams and the Ariadne Labs Better Birth team. We also acknowledge hospitals, nurses, and other government stakeholders who supported this study. Finally, we would like to acknowledge patients and families who gave their time to be surveyed.

## Appendix

**Table A:** Neonatal Mortality Rates and risk ratios, Model 1: adjusted for state and hospital-level clustering and Model 2: State specific mortality models, adjusted for hospital-level clustering

| Indicator | Adj Pre*<br>N=33599 |  | Adj CP <sup>1</sup><br>N=60078 |  | Adj Risk Ratio | 95% CI |
| --- | --- | --- | --- | --- | --- | --- |
|  | Deaths<br>(n) | NMR<br>Estimate | Deaths<br>(n) | NMR<br>Estimate |  |  |
| Model 1: Model Adjusted for State and hospital level-clustering |  |  |  |  |  |  |
| Neonatal Mortality<br>Rate<br>(per 1000 live<br>births) | 1386 | 50.5 | 2021 | 41.3 | 0.93 | 0.82, 1.06 |
| Model 2: Model by state, adjusted for hospital-level clustering |  |  |  |  |  |  |
| Punjab: |  |  |  |  |  |  |
| Neonatal Mortality<br>Rate<br>(per 1000 live<br>births) | 70 | 29.6 | 606 | 31.2 | 1.06 | 0.84, 1.33 |
| Karnataka |  |  |  |  |  |  |
| Neonatal Mortality<br>Rate<br>(per 1000 live<br>births) | 341 | 26.1 | 468 | 22.9 | 0.88 | 0.69, 1.12 |
| Maharashtra: |  |  |  |  |  |  |
| Neonatal Mortality<br>Rate<br>(per 1000 live<br>births) | 30 | 45.8 | 192 | 33.2 | 0.73 | 0.49, 1.08 |
| Madhya Pradesh: |  |  |  |  |  |  |
| Neonatal Mortality<br>Rate<br>(per 1000 live<br>births) | 945 | 53.9 | 755 | 52.2 | 0.97 | 0.83, 1.13 |

The two models presented in the table are:

Model 1: Model with neonatal mortality as outcome and intervention group as primary independent variable and adjusted for state. The variability of estimates was adjusted for hospital-level clustering. This is reported above

Model 2: State-specific models with neonatal mortality as the outcome and intervention group as a primary independent variable. The variability of estimates was adjusted for hospital-level clustering within a state.

**Table B:** Neonatal Mortality rate across Karnataka, Punjab, Madhya Pradesh (MP), and Maharashtra reported from NFHS-4 and -5, against CCP associated rates

| States | NFHS-4<br>(2015-2016) | NFHS-5<br>(2019-2021) | CCP associated NMR Pre-launch (2018-2020) | CCP associated NMR Post-launch (2018-2020) |
| --- | --- | --- | --- | --- |
| Karnataka | 18.5 | 15.8 | 26.1 | 22.9 |
| Punjab | 21.2 | 21.8 | 29.6 | 31.2 |
| MP | 36.9 | 29 | 53.9 | 52.2 |
| Maharashtra | 16.2 | 16.5 | 45.8 | 33.2 |

**Table C:** Pre and post data collection dates for each state

| State | Pre data Collection Start Date | Post data Collection Start Date |
| --- | --- | --- |
| Karnataka | 11th Oct 2018 | 26th Mar 2019 |
| Madhya Pradesh | 21st Nov 2018 | 24th July 2019 |
| Punjab | 10th Oct 2018 | 31st Oct 2018 |
| Maharashtra | 20th Mar 2019 | 29th Apr 2019 |

The unadjusted and hospital-level clustering adjusted risk ratio for maternal mortality per 100,000 live births, in the Post versus Pre group was 0.90 (95% CI: 0.53, 1.51) and 0.87 (95% CI:0.53, 1.43) respectively (Table D). Maternal deaths during pregnancy, during hospital delivery before discharge, abortions, home deliveries, or those who died after 28 days and before 42 days are not included

**Table D:**
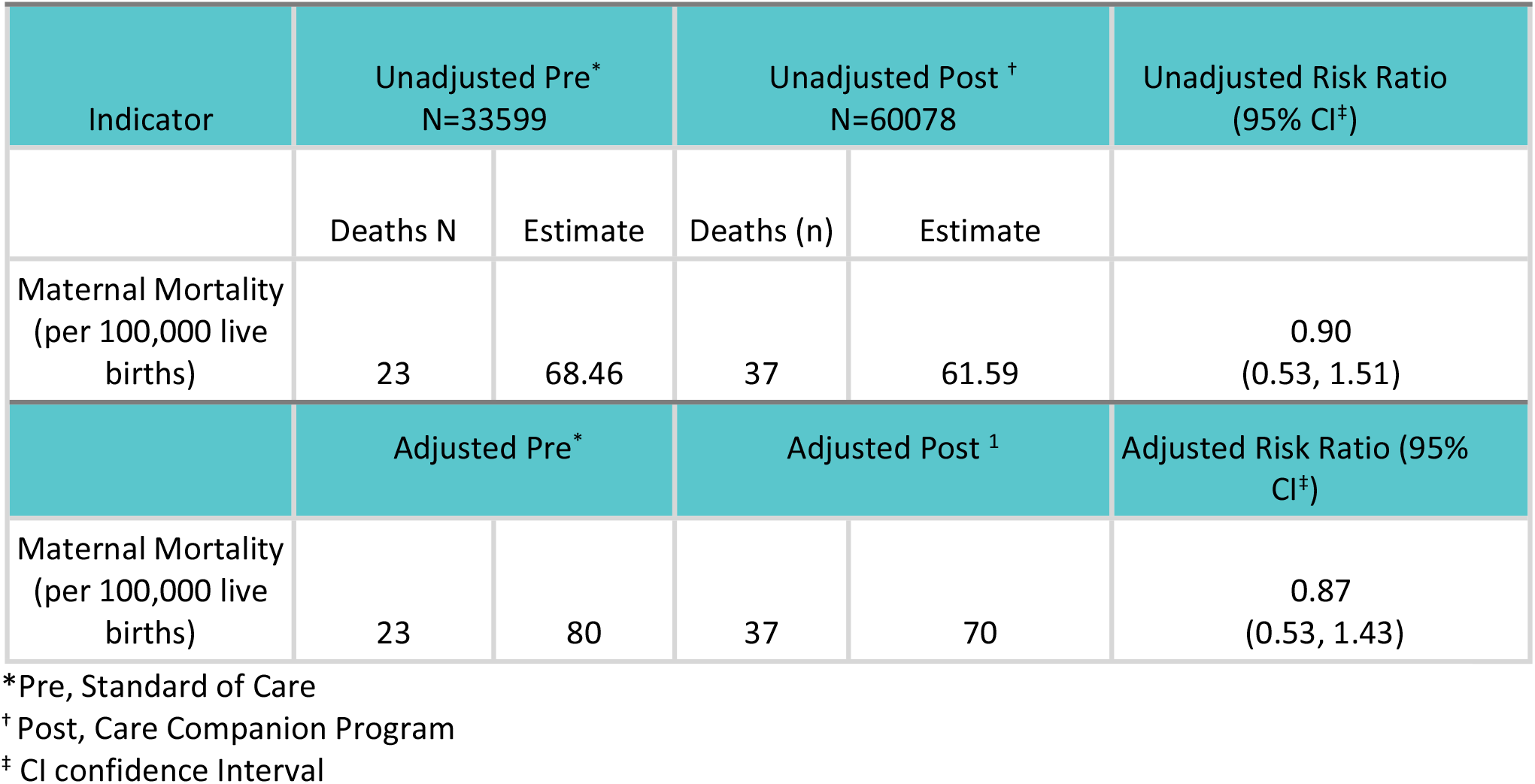
Unadjusted and hospital-level cluster adjusted estimates of Maternal Mortality and risk of mortality in Post versus Pre group.

